# ChooseMyStat: A Web-Based Interactive Tool for Statistical Test Selection and Analysis Plan Generation in Clinical Research

**DOI:** 10.64898/2026.06.02.26354730

**Authors:** Somya Srivastava, Silky R. Punyani, Deeksha Vazalwar, Ankur Joshi, Abhijit P. Pakhare

## Abstract

**Background:** Postgraduate medical residents frequently face difficulty in selecting appropriate statistical tests and preparing statistical analysis plans (SAPs) for thesis work. Existing resources often identify statistical tests without guiding implementation, reporting or software execution.

**Aims:** To describe the development, features and content validation of ChooseMyStat, a free, open-source, web-based interactive tool for statistical test selection and SAP text generation in clinical research.

**Methods:** ChooseMyStat was developed as a React-based web application using an iterative, AI-assisted development process under direct faculty supervision. The tool uses a branching decision algorithm covering 18 inferential statistical tests, two diagnostic accuracy measures, four agreement/reliability statistics, and four descriptive statistics scenarios. For each recommendation, it generates a SAP template paragraph, a results reporting example, step-by-step JASP instructions, and R code. Content validation was performed using 105 open-access original research articles from 15 broad medical specialties published in Indian journals during 2024–2025.

**Results:** The tool covers commonly used statistical methods, including t-tests, ANOVA, chi-square variants, non-parametric alternatives, correlation, regression (linear, logistic, ordinal), survival analysis, methods for clustered or repeated data, diagnostic accuracy measures, and agreement/reliability statistics. Among 365 statistical tests identified across 105 articles (excluding normality-checking procedures), 346 (94.8%) were covered by the tool. Complete coverage of all statistical methods used was observed in 86 of 105 articles (81.9%).

**Conclusions:** ChooseMyStat integrates statistical test selection with implementation guidance, SAP generation, reporting support and software instructions within a single interface. The tool may support postgraduate research training by improving accessibility to applied biostatistics guidance.

## Introduction

Selecting an appropriate statistical test is a critical yet often poorly handled aspect of clinical research, especially among postgraduate medical residents. Existing resources, whether textbook flowcharts, online decision tables, or statistical software, only name the test without guiding how to write a Statistical Analysis Plan (SAP), reporting results, or implementing the analysis in software. The ChooseMyStat tool was developed to bridge this gap by integrating test selection with Statistical Analysis Plan(SAP) generation, results reporting, and software implementation in a single, validated, freely accessible interface. This paper describes its development, features, and content validation.

## Materials and Methods

### Study design

This is a tool development and description study. Due to the lack of an appropriate reporting guideline specifically designed for educational software tools, the Template for Intervention Description and Replication (TIDieR) framework was used as a guideline for its description.^10^

### Development approach

ChooseMyStat was developed as a React-based single-page web application using Vite as the build tool and Tailwind CSS for responsive styling. The entire application runs client-side in the browser with no backend server, database, or user authentication. Development followed an iterative, AI-assisted approach: Claude (Anthropic) was used to generate initial code, decision logic drafts, SAP template text, results reporting examples, JASP instructions, and R code snippets. The senior author (AP), a faculty member in clinical epidemiology with experience supervising postgraduate theses and conducting statistical consultations, directed all design decisions and content through a collaborative iteration process—proposing requirements, reviewing AI-generated outputs, rejecting or refining recommendations, and testing each pathway against standards. All statistical content was cross-verified against established textbooks and teaching flowcharts.^5,11^

### Decision logic

The tool implements four parallel pathways: *an Inferential Test Selection Wizard, a Diagnostic Accuracy Advisor, an Agreement/Reliability Advisor, and a Descriptive Statistics Advisor*.

In the *inferential pathway*, a decision tree method is used to start the decision-making process by classifying the dependent variable based on its type (continuous, categorical/binary, ordinal, or time-to-event). The decision tree branches out further by considering other factors such as the number of groups, pairing structure, distributional assumptions (parametric vs. non-parametric), and the need for confounder adjustment. This list includes 18 different statistical tests: Independent and Paired t-tests, One-sample t-test, Mann-Whitney U, Wilcoxon signed-rank, One-way ANOVA, Kruskal-Wallis, Chi-square test of independence, Fisher’s exact test, McNemar’s test, Stuart-Maxwell test, Pearson and Spearman correlation, Linear regression, Binary logistic regression, Ordinal logistic regression, Log-rank test, Cox proportional hazards regression, Mixed-effects models, and Generalised Estimating Equations (GEE).

The *diagnostic accuracy pathway* takes users through two different scenarios: binary diagnostic test (where a two×two table is produced with sensitivity, specificity, positive and negative predictive values, and likelihood ratios) and continuous diagnostic test (ROC/AUC analysis with Youden’s Index for optimal cut-off point determination).

The *agreement/reliability pathway* encompasses four approaches depending on data type and objective: Cohen’s kappa (nominal data), Weighted kappa (ordinal data), Intraclass Correlation Coefficient (ICC; continuous reliability), and Bland-Altman analysis for comparing two continuous methods.

*Descriptive statistics* pathway involves four major cases that depend on the type of variable: continuous variables (means, medians, measures of spread), categorical variables (frequencies, proportions), ordinal variables (medians, interquartile ranges), and paired/longitudinal summaries. Each of these cases produces specific summary statistics depending on their type of variables.

### Output components

For each test suggestion, there are five output sections that are displayed in separate tabs:

#### Rationale and explanation

A clear and concise explanation of why the test selected by the tool is valid based on the inputs provided by the user, along with any assumptions and potential errors.

#### SAP template paragraph

A ready-to-adapt paragraph that can be inserted into a thesis protocol or ethics committee application, outlining the planned analysis in standard academic language. Two versions are provided: a traditional version using conventional significance testing language, and an ASA 2016-compliant version emphasising effect sizes and confidence intervals over dichotomous p-value thresholds.^12^

#### Results reporting example

An example paragraph that explains how results from the test should be stated, incorporating all the appropriate statistics, p-values (reported to three decimal places), effect sizes, and confidence intervals.

#### JASP instructions

Step-by-step instructions for running the test in JASP, a free, open-source graphical statistics package suitable for beginners.^13^

#### R code

The R code required to conduct the test, with comments explaining each step.^14^

### Analysis Plan Builder

As most theses have a number of objectives that require separate statistical procedures, the tool comes with an Analysis Plan Builder based on sessions. The user runs the selection wizard for each particular objective individually, with the results being combined by the builder in one file to download in the form of text file. It reflects the actual workflow of protocol development, where the analysis plan must map each objective to its corresponding statistical method.

### Accessibility and deployment

The application is hosted on Vercel (https://choose-my-stat.vercel.app/) (Figure.1) and is free. The application can be accessed from smartphone devices. This feature was purposefully included because many postgraduate residents from India use mobile devices as their primary means of accessing computers. The source code for the website has been made publicly available on GitHub under an open-source licence, consistent with the developers’ philosophy that a tool which reorganises existing public knowledge should itself remain openly accessible.

### Quality assurance

The validation process followed an iterative procedure. In order to ensure that the suggested test was suitable and output data is accurate, AP and AJ individually checked each decision pathway with multiple combinations of input parameters (outcome types, number of groups, paired/unpaired tests, distribution assumptions, adjustment needs) to verify that the recommended test was appropriate and the generated outputs were accurate. The suggestions were then verified against standard biostatistics references.^5,11^ SAP text, results examples, and code were assessed for accuracy, clarity, and suitability for thesis-level work. Informal peer feedback from colleagues and other postgraduate scholars was incorporated to identify potential areas of difficulty. Edge cases—such as borderline normality, small sample sizes, and scenarios requiring multiple comparison adjustments—were specifically tested and documented.

### Content validation: Coverage assessment

To assess the correspondence between the statistics covered by the tool and the methods actually used in published Indian clinical research, we conducted a cross-sectional review of open-access original research articles indexed in PubMed Central. To ensure representation across the range of all postgraduate specialties, articles were sampled from 15 broad medical disciplines: Medicine, Surgery, Paediatrics, Obstetrics & Gynaecology, Ophthalmology, Community Medicine, Psychiatry, Orthopaedics, Radiology, Anaesthesia, Dermatology, ENT, Dentistry, Pharmacology, and Family Medicine. For each specialty, articles were identified from the corresponding Indian specialty journal(s) published during 2024–2025, with a target of approximately seven articles per specialty. The search was restricted to articles mentioning “statistical analysis” in their text, to ensure that only articles with a defined analytical component were included. Articles classified as reviews, case reports, editorials, or meta-analyses were excluded.

For each article included, two reviewers (SP, DV) separately extracted all the statistical tests that have been mentioned in the Methods and Statistical Analysis sections. All of those tests were then classified as either “covered” (present in ChooseMyStat’s decision pathways) or “not covered.” In case of any differences between reviewers’ opinions, a third reviewer (AJ) made the final decision. The post-hoc correction methods (Bonferroni, Tukey, etc.) were classified as modifiers of covered tests rather than standalone methods, since ChooseMyStat’s outputs include guidance on multiple comparison adjustments where relevant. Normality assessment procedures (Kolmogorov-Smirnov, Shapiro-Wilk) identified in the articles were excluded from both the numerator and denominator of the coverage calculation, as ChooseMyStat incorporates distributional assumption as a user-entered decision parameter rather than a standalone test — consistent with its design scope. Coverage was calculated as the proportion of all identified analytical tests that fell within the tool’s scope, and the frequency of each test was tabulated.

### Ethical considerations

Since this study focuses on tool development without any involvement with any kind of human subject, there was no need to obtains formal ethics approval.

## Results

### Tool overview

ChooseMyStat is accessible at https://choosemystat.vercel.app. The home screen presents four entry points: an inferential test selection wizard, a diagnostic accuracy advisor, an agreement/reliability advisor, and a descriptive statistics advisor. A brief *How to Use* guide and a *Core Concepts* glossary are displayed on the home screen to orient first-time users. An *About* tab provides detailed explanations for each statistical test covered by the tool. [Figure 1.]

**Figure 1:**
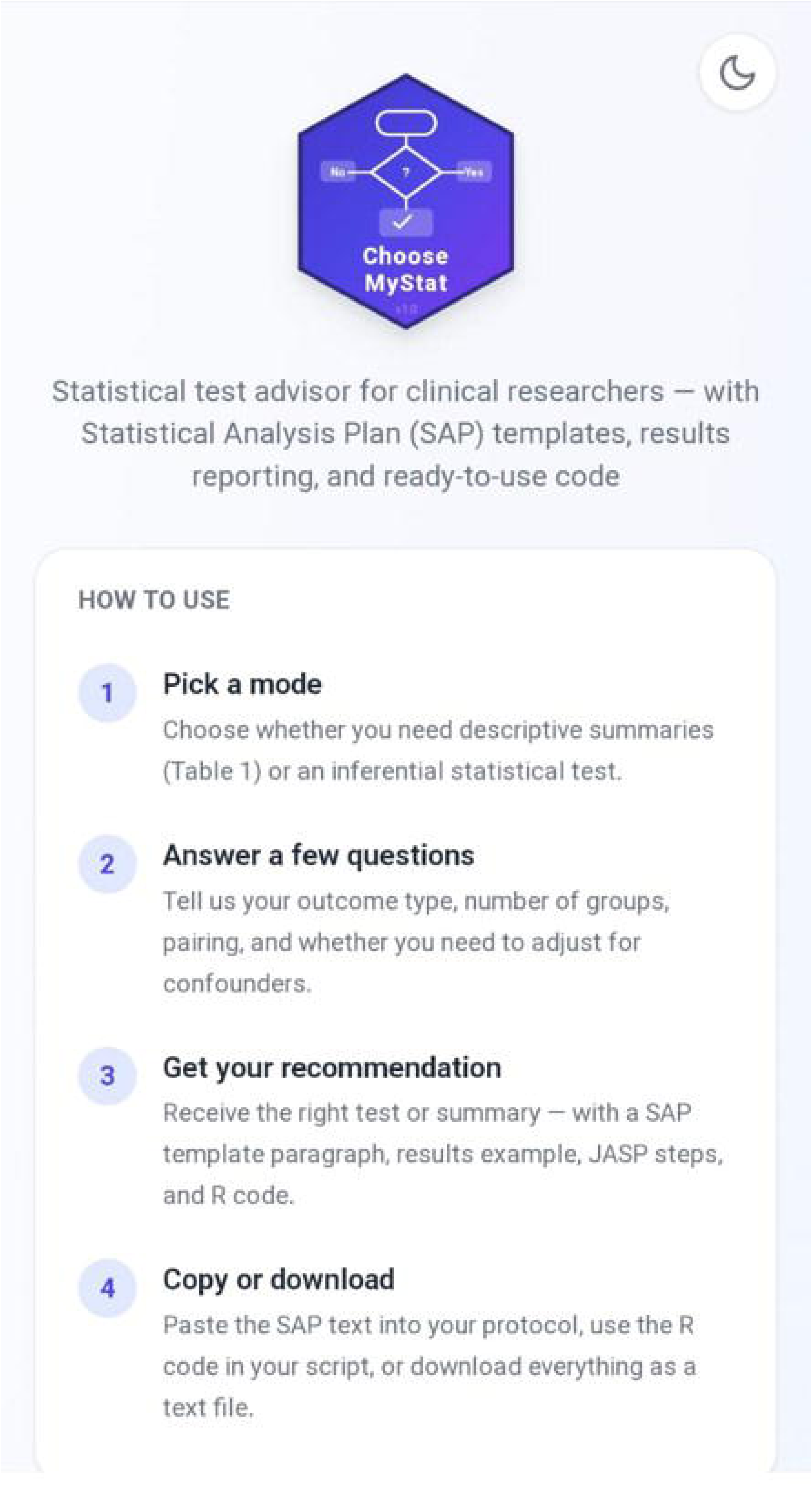
A screenshot of “Choose my stats” home screen.

### Coverage of statistical tests

Tables 1–4 present the analytical methods and decision framework implemented in the tool, including inferential tests for group comparisons (Table 1), correlation analyses (Table 2), multivariable models (Table 3), and specialised methods such as diagnostic accuracy, agreement/reliability statistics, and descriptive approaches (Table 4). In total, the tool covers 24 analytical methods along with four descriptive scenarios commonly encountered in clinical and public health research.

**Table 1:** Selection of Statistical Tests for Comparison of Groups.

| Outcome Type | Groups | Pairing | Distribution | Recommended Test |
| --- | --- | --- | --- | --- |
| Continuous | 1 | – | Normal | One-sample t-test |
| Continuous | 2 | Independent | Normal | Independent sample t-test |
| Continuous | 2 | Paired | Normal | Paired t-test |
| Continuous | 2 | Independent | Non-normal | Mann-Whitney U test |
| Continuous | 2 | Paired | Non-normal | Wilcoxon signed-rank test |
| Continuous | >2 | Independent | Normal | One-way *ANOVA |
| Continuous | >2 | Independent | Non-normal | Kruskal-Wallis test |
| Categorical | $\geq 2$ | Independent | – | Chi-square test |
| Categorical | $\geq 2$ | Independent | Small sample | Fisher's exact test |
| Categorical | 2 | Paired | – | McNemar's test |
| Categorical | >2 | Paired | – | Stuart-Maxwell test |

Note: \*ANOVA: Analysis of Variance

**Table 2:** Selection of Statistical Tests for Association and Correlation.

| Variable Type | Distribution | Recommended Test |
| --- | --- | --- |
| Continuous | Normal | Pearson correlation |
| Continuous/Ordinal | Non-normal | Spearman correlation |

**Table 3:**
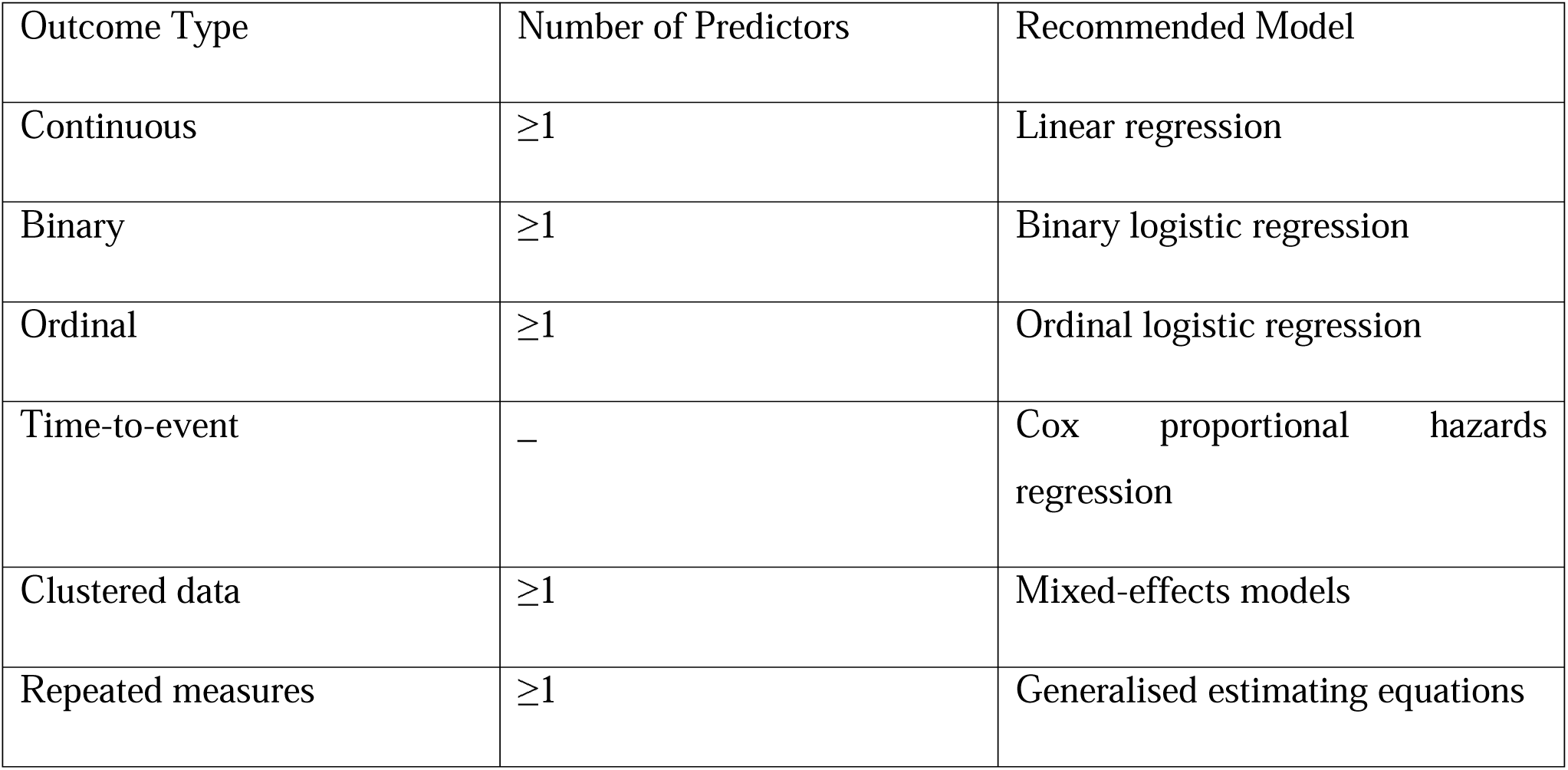
Selection of Statistical Models for Adjusted and Multivariable Analysis.

**Table 4:** Selection of Statistical Methods for Specialized Analysis.

| Diagnostic Accuracy |  |
| --- | --- |
| Data Type | Recommended Method |
| Binary outcome | Sensitivity, Specificity, <sup>*</sup>PPV, <sup>†</sup>NPV |
| Continuous outcome | <sup>‡</sup>ROC curve / <sup>§</sup>AUC |
| Agreement / Reliability |  |
| Nominal | Cohen's kappa |
| Ordinal | Weighted kappa |
| Continuous | Intraclass correlation coefficient (ICC) |
| Continuous (method comparison) | Bland-Altman analysis |
| Descriptive Statistics |  |
| Continuous | Mean ± Median <sup>¶</sup>(IQR) |
| Categorical | Frequency (%) |
| Ordinal | Median <sup>¶</sup>(IQR) |
| Longitudinal | Change scores / repeated summaries |

Note: \*PPV: Positive Predictive Value; <sup>†</sup>NPV: Negative Predictive value; <sup>‡</sup>ROC: Receiver Operating Characteristics; <sup>§</sup>AUC: Area under the curve; <sup>¶</sup>IQR: Interquartile Range.

### Workflow demonstration

To illustrate the typical workflow, consider a resident planning a study comparing systolic blood pressure between two independent groups (e.g., intervention vs. control). After selecting the inferential pathway, the resident answers four questions: (1) outcome type: continuous; (2) number of groups: two; (3) pairing: independent; (4) distribution: normally distributed. The tool recommends the independent samples t-test and immediately generates five tabbed outputs: a rationale explaining the appropriateness of the t-test given two independent groups with a normally distributed continuous outcome; a SAP template paragraph (in both traditional and ASA 2016 styles); a results reporting example with placeholder values; step-by-step JASP instructions; and R code using the *t.test()* function. (Figure 2). The entire process takes under two minutes.

**Figure 2:**
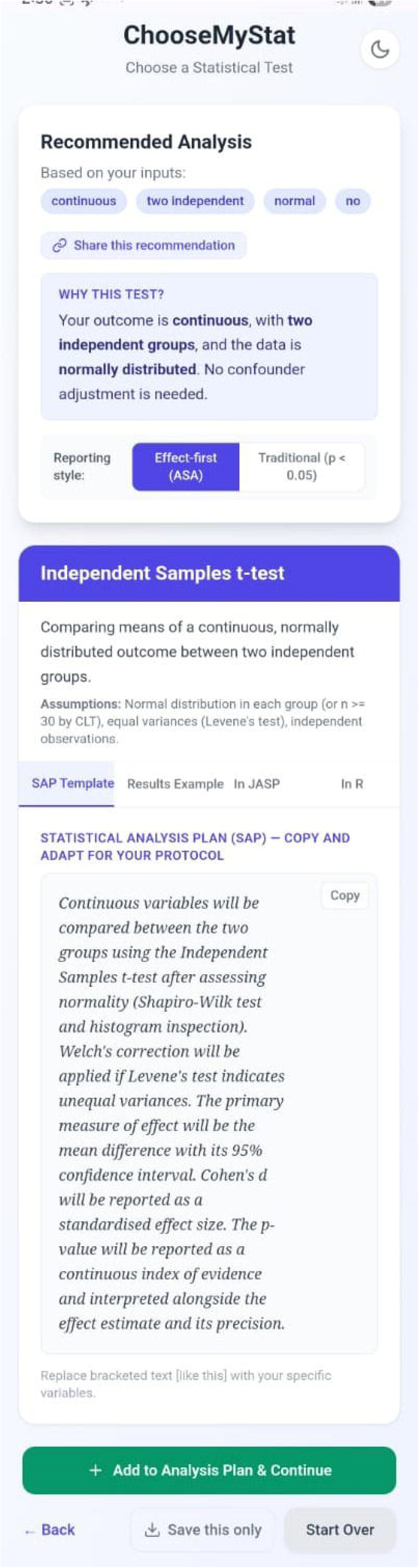
An illustrative worked example.

If the same resident indicates that the data are not normally distributed, the tool instead recommends the Mann-Whitney U test and generates correspondingly different outputs. If the resident indicates a need to adjust for confounders, the tool escalates to linear regression with appropriate modifications to all output components.

### Analysis Plan Builder

For a thesis with three objectives—(1) comparing blood pressure between groups, (2) assessing the association between a categorical exposure and a binary outcome, and (3) examining survival differences—the resident runs the wizard three times. The Analysis Plan Builder accumulates each recommendation and generates a consolidated text document mapping each objective to its statistical method, complete with SAP text and software instructions. This consolidated output can be downloaded and directly adapted for protocol submission.

### Reporting style toggle

Each output includes a toggle between traditional reporting (using conventional significance testing language such as *p < 0.05*) and ASA 2016-compliant reporting (emphasising effect sizes, confidence intervals, and exact p-values reported to three decimal places).^12^ This dual presentation serves a pedagogical purpose: residents see both styles side by side, which exposes them to evolving best practices in statistical reporting while remaining compatible with the expectations of institutional ethics committees and Indian journals that still follow traditional conventions.

### Content validation: Coverage of published research

A total of 105 open-access original research articles from 15 medical specialties, published in Indian journals during 2024–2025, were included in the validation dataset (Table 6 with complete details in Supplementary e-Table). Seven articles were sampled per specialty. Study designs included cross-sectional, cohort, randomised controlled trials, before–after studies, case–control studies, and diagnostic accuracy studies.

Across these articles, 365 analytical tests were identified (normality-checking procedures excluded). Of these, 346 tests (94.8%) were covered by ChooseMyStat. The most commonly used test was the chi-square test. The details of the tests are described in Table 5.

**Table 5:** Details of the statistical tests used in the 105 articles used for coverage validation. Normality assessment procedures (Kolmogorov-Smirnov, Shapiro-Wilk) are excluded from this table and from coverage calculations, as ChooseMyStat incorporates distributional assumption as a user-entered decision parameter.

| Statistical Test | Articles Using It (n) | % of Articles | Covered by Choose-MyStat |
| --- | --- | --- | --- |
| Chi-square test | 73 | 69.5% | Yes |
| Independent samples t-test | 53 | 50.5% | Yes |
| Paired t-test | 26 | 24.8% | Yes |
| Mann-Whitney U test | 34 | 32.4% | Yes |
| Pearson correlation | 19 | 18.1% | Yes |
| Fisher's exact test | 22 | 21.0% | Yes |
| One-way ANOVA | 16 | 15.2% | Yes |
| Diagnostic accuracy (2×2) | 10 | 9.5% | Yes |
| Kruskal-Wallis test | 17 | 16.2% | Yes |
| Binary logistic regression | 14 | 13.3% | Yes |
| Spearman correlation | 11 | 10.5% | Yes |
| ROC/AUC analysis | 11 | 10.5% | Yes |
| Linear regression | 10 | 9.5% | Yes |
| Cox proportional hazards | 5 | 4.8% | Yes |
| Log-rank test | 6 | 5.7% | Yes |
| Wilcoxon signed-rank test | 6 | 5.7% | Yes |
| Repeated measures ANOVA | 4 | 3.8% | No |
| Cohen's kappa | 1 | 1.0% | Yes |
| ICC | 2 | 1.9% | Yes |
| Poisson regression | 2 | 1.9% | No |
| Z-test/proportion test | 1 | 1.0% | No |
| McNemar's test | 2 | 1.9% | Yes |
| One-sample t-test | 1 | 1.0% | Yes |
| GEE | 1 | 1.0% | Yes |
| Mixed-effects model | 1 | 1.0% | Yes |
| Weighted kappa | 1 | 1.0% | Yes |
| Friedman test | 1 | 1.0% | No |
| Structural equation modelling | 1 | 1.0% | No |
| Factor analysis/PCA | 1 | 1.0% | No |
| Kendall's Tau Correction | 1 | 1.0% | No |

**Table 6:** Content validation results showing coverage of identified statistical tests across medical specialities.

| Specialty | Articles (n) | Tests Identified | Tests Covered | Coverage (%) | Articles fully covered |
| --- | --- | --- | --- | --- | --- |
| Medicine | 7 | 30 | 30 | 100.0% | 7 |
| Surgery | 7 | 20 | 17 | 85.0% | 4 |
| Paediatrics | 7 | 28 | 27 | 96.4% | 6 |
| Obstetrics & Gynaecology | 7 | 32 | 29 | 90.6% | 4 |
| Ophthalmology | 7 | 27 | 26 | 92.6% | 5 |
| Community Medicine | 7 | 17 | 16 | 94.1% | 6 |
| Psychiatry | 7 | 22 | 20 | 90.9% | 5 |
| Orthopaedics | 7 | 34 | 34 | 100.0% | 7 |
| Radiology | 7 | 32 | 32 | 100.0% | 7 |
| Anaesthesia | 7 | 27 | 26 | 96.3% | 6 |
| Dermatology | 7 | 23 | 21 | 91.3% | 5 |
| *ENT | 7 | 19 | 19 | 100.0% | 7 |
| Dentistry | 7 | 24 | 23 | 95.8% | 6 |
| Pharmacology | 7 | 19 | 16 | 84.2% | 4 |
| Family<br>Medicine | 7 | 12 | 12 | 100.0% | 7 |
| Total | 105 | 365 | 346 | 94.8% | 86 |

Note: \*ENT: Ear, Nose and Throat

A small number of statistical methods were not covered, primarily representing specialised techniques such as meta-analytical methods, repeated measures ANOVA, Poisson regression, and advanced multivariate approaches. These methods fall outside the intended scope of routine thesis-level analyses.

Overall, 86 of 105 articles (81.9%) had complete coverage of all statistical tests used. Coverage across specialties remained consistently high, ranging from 84.2% (Pharmacology) to 100% in multiple specialties. In the remaining articles, uncovered methods typically represented supplementary or advanced analyses rather than primary statistical approaches.

### Descriptive statistics pathway

The descriptive pathway guides users through characterising their study sample for a “Table 1” presentation. Based on variable type (continuous, categorical, ordinal, or paired/longitudinal), it recommends appropriate summary statistics (means with standard deviations, medians with interquartile ranges, frequencies with percentages) and provides formatting examples, SAP text, and software code.

## Discussion

ChooseMyStat addresses a gap in existing resources by bridging statistical test selection with implementation in a thesis protocol. Most available tools and flowcharts stop at naming the test, but that is only one part of the problem. Once a test is selected, residents struggle to explain why it is appropriate, write the analysis plan into their protocol, report results correctly, and execute the analysis in software. ChooseMyStat integrates all these steps: a user enters basic information about their research design and receives the appropriate test, SAP text, a results reporting example, step-by-step JASP instructions, and R code.

The SAP generation feature deserves particular emphasis because the analysis section is the part of the protocol that most frequently requires revision. Residents typically either leave it vague or copy text from a senior’s thesis that may not match their own design. By generating SAP text specific to the user’s chosen test and linked to a transparent decision pathway, ChooseMyStat helps residents produce a more accurate and defensible analysis plan on the first attempt, potentially saving revision cycles before ethics committee approval.

### Pedagogical Design Decisions

The question-driven wizard format was a deliberate pedagogical choice. A decision tree with 18 endpoints, presented as a complete flowchart, can overwhelm beginners. Displaying one decision at a time simplifies the learning task.^15^ Most students do not successfully navigate a full flowchart anyway — they get stuck halfway through. This step-by-step format mirrors how thesis work actually happens: solving the immediate analytical problem rather than learning all of statistics at once. With repeated use, users begin to recognise decision patterns and build understanding through application rather than memorisation.

The inclusion of both traditional and ASA 2016-compliant reporting styles reflects a pragmatic stance. While the ASA’s 2016 statement on statistical significance is reasonable,^12^ many ethics committees, thesis evaluation panels, and Indian journals continue to expect traditional reporting. Rather than imposing one approach, the toggle exposes users to both, illustrating what is changing in statistical reporting practice — balancing ideal recommendations with real-world requirements.

Offering both JASP and R instructions follows the same reasoning. JASP’s point-and-click interface suits beginners without coding experience,^13^ while R offers greater flexibility and is an industry standard for reproducibility.^14^ Given that departments and guides have varying software preferences, offering both best supports learners at different levels.

### Comparison with Existing Tools

The UCLA Institute for Digital Research and Education’s What Statistical Test Should I Use? page is among the most commonly cited online resources for test selection.^6^ It offers a comprehensive table mapping data characteristics to appropriate tests, but it is static and requires users to interpret where their study fits — difficult for those unfamiliar with the concepts. It does not generate SAP text, provide software instructions, or cover descriptive statistics.

StatsToDo and GraphPad’s test picker offer interactive guidance for quick test identification,^7^ but go no further — they do not help format the analysis plan, report results, or run the test in software. These are precisely the areas where postgraduate residents in India, learning informally under significant time constraints, encounter the greatest difficulty.

AI chatbots are increasingly used by residents^9^ but generate responses probabilistically, potentially giving different recommendations for the same scenario depending on phrasing. ChooseMyStat follows a deterministic, expert-verified decision pathway: the same inputs always produce the same output. This consistency means the tool can be audited, taught against, and systematically improved — something a chatbot’s output cannot offer.

The most immediate measurable impact of adoption in postgraduate programmes would be fewer statistical errors at the protocol stage and reduced revision cycles before ethics committee approval, since the tool directly targets the most common source of protocol revision: an unclear or incorrect analysis plan. Residents who work through the decision process may also be better positioned to justify their analytical choices during viva examinations and academic presentations. ChooseMyStat is not a substitute for a biostatistician, complex studies will always require expert consultation but it certainly resolves the routine tasks. Integration into the curriculum could take the form of a pre-thesis orientation and self-directed use during protocol development.

### AI-Assisted Development: Transparency and Epistemological Considerations

AI was used solely for application development — generating code, drafting content, and organising information — with no role in statistical decision-making. Every decision point, recommendation, and output template was reviewed by the authors against established biostatistics principles and their own teaching and consultation experience.

### Strengths and Limitations

The key advantage of ChooseMyStat is the integration of decision-making, explanation, and implementation into a single workflow. The content validation study, analysing 365 tests across 15 medical specialties, showed 94.8% coverage. The tool is free, requires no installation or registration, runs in the browser, and is mobile-responsive—all important for the intended audience.

The primary limitation is the absence of a user-level evaluation. The content validation confirms coverage of published methods, but the tool has not been assessed in a controlled study measuring its impact on test selection accuracy, SAP quality, or protocol revision rates. Such a study is a natural next step.

The scope does not cover meta-analytical methods, Bayesian approaches, repeated measures ANOVA, or complex study designs such as factorial, crossover, or cluster-randomised trials. These represent less than 5% of tests in published Indian clinical research and can be added in future versions. The absence of a backend limits reporting of feature-level engagement data, though this was a deliberate choice to prioritise simplicity and user privacy. Finally, as with any guided tool, the risk of mechanical use without understanding cannot be eliminated, though the rationale tab addresses this partially.

### Conclusion

ChooseMyStat is a practical, freely accessible tool that helps clinical researchers, particularly postgraduate medical residents to move from confusion about statistical test selection to a structured, defensible analysis plan. A content validation study across 15 medical specialties confirms that it covers 94.8% of statistical methods used in published Indian clinical research. We recommend that departments and training programmes consider incorporating such tools into their research methodology curriculum, not as replacements for teaching but as a practical tool available at the point of need.

## Data Availability

All data produced in the present work are contained in the manuscript

## Contribution Details

**Conceptualisation:** AP, AJ

**Methodology:** AP, AJ

**Software development:** AP (AI-assisted, under direct supervision)

**Validation (tool pathways):** AP, AJ

**Data acquisition (content validation):** SP, DV

**Data analysis (content validation):** AJ (independent verification)

**Statistical analysis:** AP, AJ

**Manuscript preparation:** SS, SP, AP

**Manuscript editing:** All authors

**Manuscript review:** All authors

## Guarantor

Abhijit Pakhare (AP) takes responsibility for the integrity of the work as a whole, from inception to published article.

## Acknowledgments

The ChooseMyStat web application was developed using a reproducible React-based pipeline. AI (Claude, Anthropic) was used to: (1) generate application code and user interface components under the senior author’s (AP) direct supervision and iterative review; (2) draft initial versions of the decision logic, SAP templates, results reporting examples, JASP instructions, and R code, all of which were independently reviewed and refined by AP and AJ against standard biostatistics references; and (3) assemble the present manuscript from co-author micro-question responses, preserving each author’s language and interpretive voice. All design decisions, statistical content, validation, and intellectual substance are the responsibility of the named authors. The micro-question methodology ensured that the final manuscript text reflects human expertise and authorial voice rather than AI-generated prose.

## Authorship Criteria and Approval Statement

All authors have made substantial contributions to (a) the concept and design of the study and/or acquisition, analysis, and interpretation of data; (b) drafting the article or revising it critically for important intellectual content; and (c) final approval of the version to be published. The manuscript has been read and approved by all named authors, the requirements for authorship have been met, and each author believes the manuscript represents honest work.

## Generative AI Disclosure

Claude (Anthropic) was used in the development of the ChooseMyStat application (code generation, content drafting) and in the assembly of this manuscript. The specific uses are described in full in the Acknowledgments section and the Methods section (Development approach) of the manuscript. All authors take full responsibility for the content of the manuscript.

## Clinical Trial Registration

Not applicable. This is a tool development and description study with no human subject experimentation.

## Previous Publication / Presentations

This manuscript has not been previously published, presented at any conference, or submitted to any other journal simultaneously.

## Statement of Originality

The manuscript is being submitted to the Journal of Postgraduate Medicine alone and has not been published anywhere, simultaneously submitted, or accepted for publication elsewhere.

## Author Contributions

Conceptualisation: AP, AJ. Methodology: AP, AJ. Software: AP (AI-assisted development under direct supervision). Validation (tool pathways): AP, AJ. Validation (content validation study): SP, DV extracted and classified statistical tests; AJ independently verified the classification. Writing — original draft: SS, SP, AP. Writing — review and editing: All authors. Supervision: AP, AJ. Project administration: AP. All authors read and approved the final manuscript.

## Data Availability

The content validation dataset (article-level extraction with test classification) is provided as the Supplementary e-Table. The ChooseMyStat source code is publicly available on GitHub under an open-source licence. https://github.com/drpakhare/ChooseMyStat. Its open-source code invites community review, contribution, and adaptation.

## Acknowledgments

The ChooseMyStat web application was developed using a reproducible React-based pipeline. AI (Claude, Anthropic) was used to: (1) generate application code and user interface components under the senior author’s (AP) direct supervision and iterative review; (2) draft initial versions of the decision logic, SAP templates, results reporting examples, JASP instructions, and R code, all of which were independently reviewed and refined by AP and AJ against standard biostatistics references; and (3) assemble the present manuscript from co-author micro-question responses, preserving each author’s language and interpretive voice. All design decisions, statistical content, validation, and intellectual substance are the responsibility of the named authors.

