## supplementary e-table for "ChooseMyStat: A Web-Based Interactive Tool for Statistical Test Selection and Analysis Plan Generation in Clinical Research"

**Supplementary e-Table: Article-Level Content Validation Dataset**

| **No.** | **Specialty** | **Authors** | **Year** | **Title** | **Journal** | **Study Design** | **PMC ID** | **Tests Covered by ChooseMyStat** | **Tests NOT Covered** | **Total** | **Covered (n)** | **Not Covered (n)** | **All Covered?** |
| --- | --- | --- | --- | --- | --- | --- | --- | --- | --- | --- | --- | --- | --- |
| 1 | Anaesthesia | Ahmed S et al. | 2024 | The effect of adding dexmedetomidine or dexamethasone to bupivacaine-fentanyl mixture in spinal anesthesia for cesarean | Journal of Anaesthesiology, Clinical Pharmacology | RCT | PMC11042101 | Mann-Whitney U test, One-way ANOVA, Kruskal-Wallis test, Chi-square test, post hoc Tukey's HSD test | - | 5 | 5 | 0 | **Yes** |
| 2 | Anaesthesia | Goswami D et al. | 2024 | To assess the analgesic efficacy of adjuvant magnesium sulfate added with ropivacaine over ropivacaine alone as a continuous infiltration in total abdominal hysterectomy wound | Journal of Anaesthesiology, Clinical Pharmacology | RCT | PMC11042103 | Independent samples t-test, Paired t-test, Mann Whitney U Test, Chi-square test, Fisher's exact test, GEE | - | 6 | 6 | 0 | **Yes** |
| 3 | Anaesthesia | Khurana H et al. | 2024 | A study on the outcome of preoperative pulmonary function tests on a patient undergoing rheumatic mitral valve surgery | Journal of Anaesthesiology, Clinical Pharmacology | Cohort | PMC11463928 | Independent samples t-test, Paired t-test, One-way ANOVA, Pearson correlation | - | 4 | 4 | 0 | **Yes** |
| 4 | Anaesthesia | Kumar A et al. | 2024 | Effect of different doses of dexmedetomidine as an adjuvant to lignocaine nebulization | Journal of Anaesthesiology, Clinical Pharmacology | Cohort | PMC11042085 | One-way ANOVA, post hoc Tukey's test | - | 2 | 2 | 0 | **Yes** |
| 5 | Anaesthesia | Puthenveettil N et al. | 2024 | Comparison of induction of spinal anesthesia in sitting position with legs parallel and crossed for cesarean section | Journal of Anaesthesiology, Clinical Pharmacology | RCT | PMC11042076 | Independent samples t-test, Chi-square test | - | 2 | 2 | 0 | **Yes** |
| 6 | Anaesthesia | Bansal P et al. | 2025 | Evaluation of abdominal expiratory muscle thickness pattern, diaphragmatic excursion, diaphragmatic thickness fraction and lung ultrasound score in critically ill patients | Journal of Anaesthesiology, Clinical Pharmacology | Cohort | PMC12002685 | One-way ANOVA, post hoc Tukey's test, Diagnostic accuracy (2x2), ROC/AUC analysis | - | 4 | 4 | 0 | **Yes** |
| 7 | Anaesthesia | Sharma K et al. | 2025 | Impact of anesthetic induction with etomidate, thiopentone, and propofol on regional cerebral oxygenation | Journal of Anaesthesiology, Clinical Pharmacology | Cohort | PMC11867366 | Kruskal-Wallis test, Chi-square test, Mixed-effects model | Repeated measures ANOVA | 4 | 3 | 1 | No |
| 8 | Community Medicine | AbdulRaheem Y | 2024 | Effectiveness of Awareness Training on Birth Preparedness and Complication Readiness among Community Health Workers | Indian Journal of Community Medicine | Before-after | PMC11633268 | Paired t-test, McNemar test | - | 1 | 1 | 0 | **Yes** |
| 9 | Community Medicine | Bellamkonda P et al. | 2024 | Traumatic Dental Injuries and their Association with Demographics and Other Predisposing Risk Factors in School Children | Indian Journal of Community Medicine | Cross-sectional | PMC11198536 | Chi-square test | - | 1 | 1 | 0 | **Yes** |
| 10 | Community Medicine | Ganapathy S et al. | 2025 | Determinants of Quality Antenatal Care in Pregnant Women Using NFHS-4 Data: A Path Analysis Approach | Indian Journal of Community Medicine | Cohort | PMC12156105 | Chi-square test, Fisher's exact test, Univariate logistic regression | Structural equation modeling | 4 | 3 | 1 | No |
| 11 | Community Medicine | Kumar R et al. | 2025 | Predictive Modelling of Low Birth Weight in Pregnancies: A Comparative Analysis of Logistic Regression and Decision Tree | Indian Journal of Community Medicine | Cross-sectional | PMC12430836 | Independent samples t-test, Paired t-test, Chi-square test, Binary logistic regression, Diagnostic accuracy (2x2), ROC/AUC analysis | - | 6 | 6 | 0 | **Yes** |
| 12 | Community Medicine | Prathyusha L et al. | 2025 | Out-of-pocket Expenditure on Treatment of Diabetes Mellitus and its Complications among Residents of Slum Areas of Agra | Indian Journal of Community Medicine | Cross-sectional | PMC12080904 | Mann-Whitney U test | - | 1 | 1 | 0 | **Yes** |
| 13 | Community Medicine | Priya S et al. | 2025 | Prevalence of Low Bone Mineral Density among Narikuravar Women of 18-45 Years of Age | Indian Journal of Community Medicine | Cross-sectional | PMC12080893 | Chi-square test, Pearson correlation | - | 2 | 2 | 0 | **Yes** |
| 14 | Community Medicine | Gothwal M et al. | 2026 | The Influence of Proximate Determinants on Fertility Awareness among Women Seeking Care at Infertility Clinic | Indian Journal of Community Medicine | Cross-sectional | PMC12981389 | Chi-square test, Pearson correlation | - | 2 | 2 | 0 | **Yes** |
| 15 | Dentistry | Alqarni A | 2024 | Analysis of total lip score system and total groove score for gender identification | Journal of Oral and Maxillofacial Pathology | Cross-sectional | PMC11329097 | Independent samples t-test, Paired t-test, Mann-Whitney U test, Chi-square test, Pearson correlation, Spearman correlation, Binary logistic regression, Weighted kappa | - | 8 | 8 | 0 | **Yes** |
| 16 | Dentistry | Benitha G et al. | 2024 | Molecular expression of Forkhead Box C2 gene (FOXC2) and Prospero homeobox gene (PROX-1) in oral squamous carcinoma | Journal of Oral and Maxillofacial Pathology | Cohort | PMC11329087 | Chi-square test, Linear regression, Log-rank test, Cox proportional hazards | Kendall's tau correlation | 6 | 5 | 1 | No |
| 17 | Dentistry | Chandavarkar V et al. | 2024 | Efficacy of nutritional supplement (Haras) on carbon monoxide levels in smokers and non-smokers | Journal of Oral and Maxillofacial Pathology | Cohort | PMC11126257 | Wilcoxon signed-rank test | - | 1 | 1 | 0 | **Yes** |
| 18 | Dentistry | Narang B et al. | 2024 | Potential of phosphatase and tensin gene polymorphisms as salivary biomarkers in oral squamous cell carcinoma | Journal of Oral and Maxillofacial Pathology | Cross-sectional | PMC11819637 | Independent samples t-test, Chi-square test | - | 2 | 2 | 0 | **Yes** |
| 19 | Dentistry | Muthumanickam P et al. | 2025 | Correlation of the WPOI and clinicopathological parameters in tongue OSCC patients | Journal of Oral and Maxillofacial Pathology | Cohort | PMC12283047 | Pearson correlation | - | 1 | 1 | 0 | **Yes** |
| 20 | Dentistry | Roy J et al. | 2025 | A comparative evaluation of light microscopic features of normal oral mucosa and its time dependent autolytic changes | Journal of Oral and Maxillofacial Pathology | Cross-sectional | PMC12829620 | Kruskal-Wallis test | - | 1 | 1 | 0 | **Yes** |
| 21 | Dentistry | Sharmila K et al. | 2025 | Volumetric analysis of maxillary canine and maxillary sinus for age estimation using CBCT scans | Journal of Oral and Maxillofacial Pathology | Cross-sectional | PMC12829614 | Mann-Whitney U test, Kruskal-Wallis test, Pearson correlation, Spearman correlation, Linear regression | - | 5 | 5 | 0 | **Yes** |
| 22 | Dermatology | Agrawal K et al. | 2024 | Evaluating Fractional CO2 Laser Versus Microneedling in Atrophic Acne Scars: A Split Face Study | Indian Dermatology Online Journal | Cohort | PMC11616935 | Independent samples t-test, Paired t-test, Mann-Whitney U test, Wilcoxon signed-rank test | - | 4 | 4 | 0 | **Yes** |
| 23 | Dermatology | Asfiya A et al. | 2025 | Trichoscopic Analysis of Healthy Indian Males for Standardization of the Measurable Parameters | Indian Dermatology Online Journal | Cross-sectional | PMC11753574 | Independent samples t-test, Paired t-test | Repeated measures ANOVA | 3 | 2 | 1 | No |
| 24 | Dermatology | Begum F et al. | 2025 | Comparison of Topical Mometasone, Calcipotriol, and Tacrolimus in Patients with Localized Alopecia Areata: A Triple-Arm RCT | Indian Dermatology Online Journal | RCT | PMC11927989 | One-way ANOVA, Kruskal-Wallis test, Chi-square test | - | 3 | 3 | 0 | **Yes** |
| 25 | Dermatology | Kanagarajan A et al. | 2025 | Low Dose Vs Conventional Rituximab Regimen in Pemphigus Vulgaris | Indian Dermatology Online Journal | Cohort | PMC12270409 | Mann-Whitney U test, Chi-square test, Fisher's exact test, Log-rank test | - | 5 | 5 | 0 | **Yes** |
| 26 | Dermatology | Malviya A et al. | 2025 | A Prospective Study Comparing 30% Mandelic Acid Peel Versus 30% Lactic Acid Peel in Periorbital Melanosis | Indian Dermatology Online Journal | Cohort | PMC12622926 | Independent samples t-test, Paired t-test, Mann-Whitney U test, Chi-square test | Z-test | 5 | 4 | 1 | No |
| 27 | Dermatology | Reddy M et al. | 2025 | Clinical and Laboratory Parameters of Metabolic Syndrome in Chronic Spontaneous Urticaria | Indian Dermatology Online Journal | Cross-sectional | PMC11753540 | Chi-square test | - | 1 | 1 | 0 | **Yes** |
| 28 | Dermatology | Bajoria S et al. | 2026 | Phototherapy with Topical Minoxidil in Vitiligo: A Randomized Control Trial | Indian Dermatology Online Journal | RCT | PMC12854552 | Chi-square test, t-test | - | 2 | 2 | 0 | **Yes** |
| 29 | ENT | Das K et al. | 2024 | Occult Metastasis: Incidence, Pattern, and Impact on Survival in Patients with Oral Cancer | Indian Journal of Otolaryngology and Head & Neck Surgery | Cohort | PMC11569288 | Chi-square test, Pearson correlation, Kaplan-Meier | - | 3 | 3 | 0 | **Yes** |
| 30 | ENT | Krishnatreya M et al. | 2024 | High Risk Human Papillomavirus Prevalence in Patients with Hypopharynx Cancer of Northeast India | Indian Journal of Otolaryngology and Head & Neck Surgery | Cross-sectional | PMC11456070 | Chi-square test | - | 1 | 1 | 0 | **Yes** |
| 31 | ENT | Ragland A et al. | 2024 | Understanding the Vestibular Apparatus: How 3D Models Can Improve Student Learning | Indian Journal of Otolaryngology and Head & Neck Surgery | Before-after | PMC11890795 | Independent samples t-test, Chi-square test | - | 2 | 2 | 0 | **Yes** |
| 32 | ENT | Revercomb L et al. | 2024 | Bibliometrics and National Institutes of Health Funding: Associated Factors in Academic Rhinology | Indian Journal of Otolaryngology and Head & Neck Surgery | Cohort | PMC11890462 | Kruskal-Wallis test, Chi-square test | - | 2 | 2 | 0 | **Yes** |
| 33 | ENT | Aftab O et al. | 2025 | Surgical Subspecialty and Parathyroidectomy Outcomes: A National Analysis | Indian Journal of Otolaryngology and Head & Neck Surgery | Cohort | PMC11985825 | Mann-Whitney U test, Chi-square test, Binary logistic regression | - | 3 | 3 | 0 | **Yes** |
| 34 | ENT | Jacob T et al. | 2025 | Outcomes of Surgical Bed Versus Tumor Margins in Trans-Oral Resection of Early Glottic Cancer | Indian Journal of Otolaryngology and Head & Neck Surgery | Cohort | PMC12103429 | Independent samples t-test, Mann-Whitney U test, Chi-square test, Fisher's exact test | - | 4 | 4 | 0 | **Yes** |
| 35 | ENT | Pavlidis P et al. | 2025 | An Examination of Tinnitus and Cochlear Functionality in Hearing Impaired and Normally Hearing Individuals | Indian Journal of Otolaryngology and Head & Neck Surgery | Before-after | PMC12297068 | Independent samples t-test, Kruskal-Wallis test | - | 3 | 3 | 0 | **Yes** |
| 36 | Family Medicine | Abdulrahman K et al. | 2025 | Awareness regarding common eye diseases among university students at a public university in Riyadh City | Journal of Family Medicine and Primary Care | Cross-sectional | PMC12858160 | Chi-square test, Linear regression | - | 2 | 2 | 0 | **Yes** |
| 37 | Family Medicine | Almogbel E | 2025 | Transforming diabetes care: The effect of insulin pump technology on quality of life in adults with type 1 diabetes | Journal of Family Medicine and Primary Care | Case-control | PMC12858152 | Independent samples t-test, Chi-square test, Logistic regression | - | 3 | 3 | 0 | **Yes** |
| 38 | Family Medicine | Charaimuriya B et al. | 2025 | Decreasing trend of nutritional anemia in anemic pregnant mothers in hospital based study in Meghalaya | Journal of Family Medicine and Primary Care | Cross-sectional | PMC12858140 | Chi-square test | - | 1 | 1 | 0 | **Yes** |
| 39 | Family Medicine | Chawale S et al. | 2025 | An epidemiological study of depression among geriatric population in Mehrauli area of South Delhi | Journal of Family Medicine and Primary Care | Cross-sectional | PMC12858158 | Chi-square test, Binary logistic regression | - | 2 | 2 | 0 | **Yes** |
| 40 | Family Medicine | Jahnavi G et al. | 2025 | Quality of care, support and treatment services in Jharkhand's Antiretroviral Therapy Center: A mixed methods study | Journal of Family Medicine and Primary Care | Cross-sectional | PMC12858128 | Chi-square test | - | 1 | 1 | 0 | **Yes** |
| 41 | Family Medicine | Khan S et al. | 2025 | Seroprevalence of SARS CoV 2 IgG in pandemic frontline healthcare workers | Journal of Family Medicine and Primary Care | Cross-sectional | PMC12858119 | Paired t-test, Chi-square test | - | 2 | 2 | 0 | **Yes** |
| 42 | Family Medicine | Khapre M et al. | 2025 | Health seeking behavior and prevalence of self-reported symptoms of reproductive tract infection among women | Journal of Family Medicine and Primary Care | Cross-sectional | PMC12858110 | Binary logistic regression | - | 1 | 1 | 0 | **Yes** |
| 43 | Medicine | Amin N et al. | 2024 | Association between Lipoprotein(a) concentration and adverse cardiac events in patients with coronary artery disease | Indian Heart Journal | Cohort | PMC11328994 | Chi-square test, t-test, Log-rank test, Cox proportional hazards | - | 4 | 4 | 0 | **Yes** |
| 44 | Medicine | Kumar D et al. | 2024 | Relationship between high sensitivity troponin I and clinical outcomes in non-ACS acute heart failure patients | Indian Heart Journal | Cohort | PMC11143518 | Independent samples t-test, Chi-square test | - | 2 | 2 | 0 | **Yes** |
| 45 | Medicine | Priyanka S, Morkar D | 2024 | AST/ALT Ratio as an indicator of functional severity in chronic heart failure with reduced LVEF | Indian Heart Journal | Cross-sectional | PMC11329059 | Mann-Whitney U test, Chi-square test, Spearman correlation, Linear regression, Binary logistic regression, Diagnostic accuracy (2x2), ROC/AUC analysis | - | 7 | 7 | 0 | **Yes** |
| 46 | Medicine | Bhandarkar A et al. | 2025 | Bone Health in Young Adults with Type 1 Diabetes Mellitus in South India | Indian Journal of Endocrinology and Metabolism | Cross-sectional | PMC11964363 | Mann-Whitney U test, Kruskal-Wallis test, Chi-square test | - | 3 | 3 | 0 | **Yes** |
| 47 | Medicine | Ray S et al. | 2025 | Drug-coated balloon in patients with in-stent restenosis: A prospective observational study | Indian Heart Journal | Cross-sectional | PMC12138078 | Independent samples t-test, Chi-square test, Fisher's exact test, Log-rank test, Cox proportional hazards | - | 5 | 5 | 0 | **Yes** |
| 48 | Medicine | Satheesh M et al. | 2025 | Comparative Study of Two Successive American Thyroid Association Risk Stratification Systems in Differentiated Thyroid Cancer | Indian Journal of Endocrinology and Metabolism | Cohort | PMC12604840 | Chi-square test, McNemar's test, Logistic regression, Diagnostic accuracy (2x2) | - | 4 | 4 | 0 | **Yes** |
| 49 | Medicine | Yetkin G et al. | 2026 | Infectious agents, protein electrophoresis and immunoglobulin levels in patients with coronary artery ectasia | Indian Heart Journal | Cohort | PMC13040902 | Independent samples t-test, Paired t-test, Chi-square test, Fisher's exact test, Binary logistic regression | - | 5 | 5 | 0 | **Yes** |
| 50 | Obstetrics & Gynaecology | Dhar S et al. | 2024 | Serum Zinc Levels in Women with Polycystic Ovarian Syndrome | Journal of Human Reproductive Sciences | Cohort | PMC11041321 | Independent samples t-test, Paired t-test, Pearson correlation, Linear regression | - | 4 | 4 | 0 | **Yes** |
| 51 | Obstetrics & Gynaecology | Kumar P et al. | 2024 | Diagnostic Utility of Various Hormones across Different Polycystic Ovary Syndrome Phenotypes | Journal of Human Reproductive Sciences | Cross-sectional | PMC11741125 | Independent samples t-test, Mann-Whitney U test, One-way ANOVA, Kruskal-Wallis test, Diagnostic accuracy (2x2), ROC/AUC analysis | - | 6 | 6 | 0 | **Yes** |
| 52 | Obstetrics & Gynaecology | Shivhare S et al. | 2024 | Does Physiological ICSI Improve Outcome in Men with Abnormal Semen Parameters: A Retrospective Cohort Study | Journal of Human Reproductive Sciences | RCT | PMC11559347 | Independent samples t-test, Paired t-test, Chi-square test, Fisher's exact test | Poisson regression | 5 | 4 | 1 | No |
| 53 | Obstetrics & Gynaecology | Yuningsih T et al. | 2024 | Utilisation of Oocyte Diameter as a Non-invasive Indicator of Oocyte and Embryo Quality | Journal of Human Reproductive Sciences | Cohort | PMC11559353 | One-way ANOVA, Kruskal-Wallis test, Chi-square test | - | 3 | 3 | 0 | **Yes** |
| 54 | Obstetrics & Gynaecology | Bhan S et al. | 2025 | Angiotensin-converting Enzyme Insertion/Deletion Polymorphism in North Indian Population and Its Correlation with Male Infertility | Journal of Human Reproductive Sciences | Case-control | PMC12527149 | Independent samples t-test, Chi-square test, Pearson correlation, Binary logistic regression | - | 4 | 4 | 0 | **Yes** |
| 55 | Obstetrics & Gynaecology | Shah N et al. | 2025 | Association of Sperm DNA Fragmentation and ICSI Outcomes in an Age-adjusted Cohort | Journal of Human Reproductive Sciences | Cohort | PMC12815425 | Kruskal-Wallis test, Chi-square test, Pearson correlation, Binary logistic regression | - | 5 | 4 | 1 | No |
| 56 | Obstetrics & Gynaecology | Singh N et al. | 2025 | Should We Be Offering ICSI to All Couples with Unexplained Infertility: A Cohort Study | Journal of Human Reproductive Sciences | Cohort | PMC12057842 | Independent samples t-test, Mann-Whitney U test, Chi-square test, Fisher's exact test | - | 5 | 4 | 1 | No |
| 57 | Ophthalmology | Bassi S et al. | 2024 | Optical coherence tomography in papilledema: A probe into the intracranial pressure correlation | Indian Journal of Ophthalmology | Cross-sectional | PMC11168544 | Chi-square test, Pearson correlation | - | 2 | 2 | 0 | **Yes** |
| 58 | Ophthalmology | Bhattacharjee K et al. | 2024 | Central and peripheral contrast sensitivity in thyroid eye disease | Indian Journal of Ophthalmology | Cross-sectional | PMC11573027 | Mann-Whitney U test, Kruskal-Wallis test, Chi-square test, Pearson correlation, Spearman correlation | - | 5 | 5 | 0 | **Yes** |
| 59 | Ophthalmology | Dutta R et al. | 2024 | Aberration change after scleral lens wear in eyes with pellucid marginal degenerations | Indian Journal of Ophthalmology | Cohort | PMC11329822 | Independent samples t-test, Pearson correlation | - | 3 | 2 | 1 | No |
| 60 | Ophthalmology | Ahmet S et al. | 2025 | The long-term effects of interface irrigation on visual outcomes and corneal aberrations in SMILE | Indian Journal of Ophthalmology | Cohort | PMC12356364 | Independent samples t-test, Paired t-test, Mann-Whitney U test, One-way ANOVA, Kruskal-Wallis test | Repeated measures ANOVA | 7 | 6 | 1 | No |
| 61 | Ophthalmology | Bassi S | 2025 | Comparison of clinical outcomes of phacoemulsification with implantation of two new aspheric IOLs: Real-world data | Indian Journal of Ophthalmology | Cohort | PMC12178399 | Independent samples t-test, Paired t-test | - | 2 | 2 | 0 | **Yes** |
| 62 | Ophthalmology | Campos P et al. | 2025 | Comparison of tolerance to induced astigmatism in pseudophakic eyes implanted with dual-technology diffractive IOL | Indian Journal of Ophthalmology | Cohort | PMC12448531 | Independent samples t-test, Paired t-test, Mann-Whitney U test, Fisher's exact test | - | 4 | 4 | 0 | **Yes** |
| 63 | Ophthalmology | Gupta S et al. | 2025 | Comparative analysis of retinal thickness between type 1 and type 2 diabetes mellitus patients | Indian Journal of Ophthalmology | Cross-sectional | PMC12659822 | Independent samples t-test, Paired t-test, Mann-Whitney U test, Chi-square test | - | 4 | 4 | 0 | **Yes** |
| 64 | Orthopaedics | Bevan A et al. | 2024 | Can a Surgical Vulnerability Score Predict Outcomes of Hip Reconstruction in Children with Severe Neuromuscular Disability | Indian Journal of Orthopaedics | Cohort | PMC11628471 | Independent samples t-test, Paired t-test, Chi-square test, Pearson correlation | - | 4 | 4 | 0 | **Yes** |
| 65 | Orthopaedics | Chawanpaiboon P et al. | 2024 | Incidence of and Factors Associated with Spontaneous Correction of Postoperative Shoulder Imbalance in AIS | Indian Journal of Orthopaedics | Before-after | PMC11775351 | Linear regression, Binary logistic regression, ROC/AUC analysis, Cohen's kappa, ICC | - | 5 | 5 | 0 | **Yes** |
| 66 | Orthopaedics | Li S et al. | 2024 | Comparison of mMO-TLIF via Midline Incision Versus MIS-TLIF via Wiltse Approach in Lumbar Degenerative Disease | Indian Journal of Orthopaedics | RCT | PMC11333641 | Independent samples t-test, Paired t-test, Mann-Whitney U test, Kruskal-Wallis test, Chi-square test | - | 5 | 5 | 0 | **Yes** |
| 67 | Orthopaedics | Patel P et al. | 2024 | The Effect of Prone and Supine Limb Positioning on the Radiographic Evaluation of Posterolateral Plate Fixation | Indian Journal of Orthopaedics | Before-after | PMC10899138 | Mann-Whitney U test | - | 1 | 1 | 0 | **Yes** |
| 68 | Orthopaedics | Wang Z et al. | 2024 | Comparison of GAP Score and SRS-Schwab ASD Classification in Surgical Outcomes for Adult Spinal Deformity | Indian Journal of Orthopaedics | Cohort | PMC11130083 | Independent samples t-test, Paired t-test, One-way ANOVA, Chi-square test, Fisher's exact test, Pearson correlation, ROC/AUC analysis | - | 8 | 8 | 0 | **Yes** |
| 69 | Orthopaedics | Luo X et al. | 2025 | MRI Findings of Extradural Fat in Patients with Cauda Equina Syndrome: A Novel Perspective | Indian Journal of Orthopaedics | Cohort | PMC12496402 | Independent samples t-test, Wilcoxon signed-rank test, One-way ANOVA, Kruskal-Wallis test, Chi-square test, Fisher's exact test, Spearman correlation | - | 7 | 7 | 0 | **Yes** |
| 70 | Orthopaedics | Smimmo A et al. | 2025 | Clinic and Ultrasound Evaluation of Suction Drainage in Total Knee Arthroplasty Procedure | Indian Journal of Orthopaedics | Cohort | PMC12615859 | Independent samples t-test, Mann-Whitney U test, One-way ANOVA, Chi-square test | - | 4 | 4 | 0 | **Yes** |
| 71 | Pediatrics | Pedaveeti M et al. | 2024 | Comparative Growth Outcomes in Very Low Birth Weight Infants: Evaluating Different Feeding Strategies | Indian Journal of Pediatrics | RCT | PMC11913907 | Independent samples t-test, Paired t-test, One-way ANOVA, Chi-square test, Fisher's exact test | - | 5 | 5 | 0 | **Yes** |
| 72 | Pediatrics | Wang Q et al. | 2024 | Diagnostic Value of Single LH and LH/FSH Ratio at 60-minute after GnRHa Stimulation Test for Central Precocious Puberty | Indian Journal of Pediatrics | Cohort | PMC12279564 | Mann-Whitney U test, Chi-square test, Spearman correlation, Diagnostic accuracy (2x2), ROC/AUC analysis | - | 6 | 6 | 0 | **Yes** |
| 73 | Pediatrics | Altay D et al. | 2025 | Helicobacter pylori in Turkish Children with Dyspepsia: Diagnosis, Prevalence, Genotyping and Antibiotic Resistance | Indian Journal of Pediatrics | Not specified | PMC12764497 | Independent samples t-test, One-sample t-test, Mann-Whitney U test, Chi-square test | - | 5 | 4 | 1 | No |
| 74 | Pediatrics | Bafunyembaka G et al. | 2025 | Association of Asthma with Acute Vaso-Occlusive Crisis Among French Guianese Children with Sickle Cell Disease | Indian Journal of Pediatrics | Cohort | PMC12855333 | Linear regression | - | 1 | 1 | 0 | **Yes** |
| 75 | Pediatrics | Chaudhary V et al. | 2025 | Epidemiological and Clinical Characteristics of Intussusception in Children Aged Under 2 Years | Indian Journal of Pediatrics | Before-after | PMC12801410 | Chi-square test, Fisher's exact test, Rank sum test | - | 3 | 3 | 0 | **Yes** |
| 76 | Pediatrics | Yesiltepe E et al. | 2025 | FCGR2A Gene Polymorphism Association in Children with Multisystem Inflammatory Syndrome | Indian Pediatrics | Case-control | PMC12041097 | Mann-Whitney U test, Chi-square test, Fisher's exact test | - | 3 | 3 | 0 | **Yes** |
| 77 | Pediatrics | Zhong H et al. | 2025 | Effect of Macrolide Resistance and Mycoplasma pneumoniae DNA Load on Immune and Inflammatory Responses in Children | Indian Pediatrics | Cohort | PMC12644151 | Independent samples t-test, Paired t-test, Mann-Whitney U test, Chi-square test, Spearman correlation | - | 5 | 5 | 0 | **Yes** |
| 78 | Pharmacology | Agrawal A et al. | 2025 | Efficacy and safety of tranexamic acid nebulization to control bleeding of hemoptysis: The TXA-NEB RCT | Indian Journal of Pharmacology | RCT | PMC12662606 | Independent samples t-test, Paired t-test, Chi-square test | - | 4 | 3 | 1 | No |
| 79 | Pharmacology | Jhaj R et al. | 2025 | An analysis of awareness and acceptability of the Medicines Side Effect Reporting Form for Consumer | Indian Journal of Pharmacology | Cross-sectional | PMC12662611 | Independent samples t-test, Chi-square test | - | 3 | 3 | 0 | **Yes** |
| 80 | Pharmacology | Lakkanna A et al. | 2025 | Impact of the timing of antibiotic prophylaxis on surgical site infections in patients undergoing elective general surgery | Indian Journal of Pharmacology | Cohort | PMC12419558 | Independent samples t-test, Chi-square test | - | 2 | 2 | 0 | **Yes** |
| 81 | Pharmacology | Liu C et al. | 2025 | Effect of dexmedetomidine combined with ropivacaine in ultrasound-guided paravertebral nerve block for herpes zoster neuralgia | Indian Journal of Pharmacology | Cross-sectional | PMC12662612 | t-tests, Mann-Whitney U test, Chi-square test | Repeated measures ANOVA | 4 | 3 | 1 | No |
| 82 | Pharmacology | Panda S et al. | 2025 | Efficacy of epigallocatechin-3-gallate against oleic acid-induced acute respiratory distress syndrome | Indian Journal of Pharmacology | Before-after | PMC12662607 | One-way ANOVA, post hoc Tukey's test | - | 2 | 2 | 0 | **Yes** |
| 83 | Pharmacology | Tavethia J et al. | 2025 | Assessment of knowledge, attitude, and practice regarding reporting of adverse events due to medical devices | Indian Journal of Pharmacology | Cross-sectional | PMC12662616 | Independent samples t-test, Chi-square test | - | 2 | 2 | 0 | **Yes** |
| 84 | Pharmacology | Yin Y et al. | 2025 | Safety assessment of dexmedetomidine: Real-world adverse event analysis from FAERS public dashboard | Indian Journal of Pharmacology | Before-after | PMC12419569 | Chi-square test | Poisson regression | 2 | 1 | 1 | No |
| 85 | Psychiatry | Akash N et al. | 2024 | Autistic features in patients with intellectual disability attending psychiatry outpatient department | Industrial Psychiatry Journal | Cross-sectional | PMC11553612 | Chi-square test, Spearman correlation | - | 2 | 2 | 0 | **Yes** |
| 86 | Psychiatry | Chopra G, Gaur V | 2024 | Depression, anxiety, and quality of life among kidney donors before and after kidney donation | Industrial Psychiatry Journal | Cohort | PMC11784696 | Wilcoxon signed-rank test, Pearson correlation, Linear regression | - | 3 | 3 | 0 | **Yes** |
| 87 | Psychiatry | Madhusudhan G et al. | 2024 | Correlation of severity of alcohol dependence with liver dysfunction by transient elastography | Industrial Psychiatry Journal | Cross-sectional | PMC11784701 | Paired t-test, Spearman correlation | - | 2 | 2 | 0 | **Yes** |
| 88 | Psychiatry | Murugan Y et al. | 2024 | Exploring the association between depression and diabetes among type 1 and type 2 diabetic mellitus patients | Industrial Psychiatry Journal | Cross-sectional | PMC11155637 | Chi-square test, Binary logistic regression | - | 2 | 2 | 0 | **Yes** |
| 89 | Psychiatry | Gurjar N et al. | 2025 | Effects of electroconvulsive therapy on serum BDNF, interleukin-6, and cortisol in treatment refractory schizophrenia | Industrial Psychiatry Journal | Cohort | PMC12077616 | Paired t-test, Spearman correlation | - | 2 | 2 | 0 | **Yes** |
| 90 | Psychiatry | Sandhu S et al. | 2025 | A cross-sectional study on self-esteem and body image dissatisfaction as mediators in the relationship between perfectionism and suicidality | Industrial Psychiatry Journal | Cross-sectional | PMC12574771 | Independent samples t-test, Pearson correlation, Linear regression | Mediation Analysis | 4 | 3 | 1 | No |
| 91 | Psychiatry | Jindal M et al. | 2026 | Second Victim Syndrome in Indian Surgeons - A Cross Sectional Analytical Study | Industrial Psychiatry Journal | Cross-sectional | PMC12923233 | Wilcoxon signed-rank test, One-way ANOVA, Kruskal-Wallis test, Chi-square test, Spearman correlation, Linear regression | Factor analysis | 7 | 6 | 1 | No |
| 92 | Radiology | Gaurav G et al. | 2024 | Evaluation of Benign and Malignant Cervical Lymphadenopathy: Shear Wave Elastography and Grayscale Ultrasound | The Indian Journal of Radiology & Imaging | Cross-sectional | PMC12169923 | Fisher's exact test, Diagnostic accuracy (2x2), ROC/AUC analysis, ICC | - | 4 | 4 | 0 | **Yes** |
| 93 | Radiology | Halmandge A et al. | 2024 | Comparison of MRI Osteoarthritis Knee Score with Clinico-Radiological Grading | The Indian Journal of Radiology & Imaging | Cross-sectional | PMC11651822 | Descriptive analysis (frequency, mean, and median) | - | 3 | 3 | 0 | **Yes** |
| 94 | Radiology | Narayan S et al. | 2024 | Prevalence of Osteoporosis and Sarcopenia in Middle-Aged Subjects with Low Back Pain | The Indian Journal of Radiology & Imaging | Cross-sectional | PMC11651826 | One-way ANOVA, Chi-square test, t-test, Pearson correlation | - | 4 | 4 | 0 | **Yes** |
| 95 | Radiology | Chandola S et al. | 2025 | CT-Based Texture Analysis in Indeterminate Pediatric Renal and Pararenal Masses | The Indian Journal of Radiology & Imaging | Cohort | PMC12788926 | Mann-Whitney U test, Chi-square test, Fisher's exact test, Diagnostic accuracy (2x2), ROC/AUC analysis, t-test | - | 6 | 6 | 0 | **Yes** |
| 96 | Radiology | Dharmalingam P, Jagannathan D | 2025 | Comparative Analysis of Radiation Dose in Advanced X-Ray Mammographic Modalities | The Indian Journal of Radiology & Imaging | Cohort | PMC13002273 | Independent samples t-test, One-way ANOVA with post hoc corrections | - | 2 | 2 | 0 | **Yes** |
| 97 | Radiology | Singh A et al. | 2025 | Evaluation of Maternal Ophthalmic Artery Doppler Velocimetry at 18-24 Weeks in Prediction of Pre-eclampsia | The Indian Journal of Radiology & Imaging | Cohort | PMC13002269 | Mann-Whitney U test, Chi-square test, Fisher's exact test, Diagnostic accuracy (2x2), ROC/AUC analysis | - | 5 | 5 | 0 | **Yes** |
| 98 | Radiology | Thomas A et al. | 2025 | Prediction of Survival in Surgically Treated Glioblastoma Multiforme Utilizing DTI and Contrast-Enhanced MRI | The Indian Journal of Radiology & Imaging | Cohort | PMC12788931 | Mann-Whitney U test, Chi-square test, t-test, Pearson correlation, Log-rank test, Cox proportional hazards, Diagnostic accuracy (2x2), ROC/AUC analysis | - | 8 | 8 | 0 | **Yes** |
| 99 | Surgery | Ojha S et al. | 2024 | Laparoscopic choledochal cyst excision and biliary reconstruction in patients with previous surgery/intervention | Journal of Minimal Access Surgery | Cohort | PMC11095812 | Independent samples t-test, Paired t-test, Fisher's exact test | - | 3 | 3 | 0 | **Yes** |
| 100 | Surgery | Aragone L et al. | 2025 | Self-gripping mesh in laparoscopic inguinal hernia repair: A comparative study | Journal of Minimal Access Surgery | Cohort | PMC12327777 | Mann-Whitney U test, Fisher's exact test | - | 2 | 2 | 0 | **Yes** |
| 101 | Surgery | Kar S et al. | 2025 | Laparoscopy-guided transverse abdominis plane block versus port site infiltration for post-operative pain relief | Journal of Minimal Access Surgery | RCT | PMC12054959 | Mann-Whitney U test, Chi-square test, Fisher's exact test, Wilcoxon signed-rank test | Friedman test | 5 | 4 | 1 | No |
| 102 | Surgery | Meena S et al. | 2025 | Effect of scrotal support application on seroma formation following minimal access surgery for inguinal hernia | Journal of Minimal Access Surgery | RCT | PMC12054949 | Chi-square test or Fisher's exact test, Mann-Whitney U test | - | 1 | 1 | 0 | **Yes** |
| 103 | Surgery | Varun R et al. | 2025 | Telescopic dissection versus balloon dissection during laparoscopic totally extraperitoneal inguinal hernia repair | Journal of Minimal Access Surgery | RCT | PMC12054956 | Independent Student's t-test/Mann-Whitney U-test, Chi-square test/Fisher's exact | - | 3 | 2 | 1 | No |
| 104 | Surgery | Yaman S et al. | 2025 | Comparison of short-term oncologic outcomes in open, laparoscopic and robotic radical gastrectomy for gastric cancer | Journal of Minimal Access Surgery | Retrospective cross-sectional | PMC12327784 | Kruskal-Wallis test, Chi-square test | - | 4 | 3 | 1 | No |
| 105 | Surgery | Wang L, Zhou X | 2026 | Outcomes of laparoscopic inguinal hernia repair using tail-anchor mesh fixation method: A retrospective study | Journal of Minimal Access Surgery | Before-after | PMC12904628 | Independent samples t-test, Chi-square test | - | 2 | 2 | 0 | **Yes** |
| **TOTAL** | **105 articles** |  |  |  |  |  |  |  |  | **365** | **346** | **19** |  |

*Abbreviations: RCT = Randomised Controlled Trial; PMC ID = PubMed Central Identifier; GEE = Generalised Estimating Equations; ICC = Intraclass Correlation Coefficient; ROC/AUC = Receiver Operating Characteristic/Area Under the Curve.*

Summary:

| Total analytical tests identified | **365** |
| --- | --- |
| Tests covered by ChooseMyStat | **346** |
| Tests NOT covered | **19** |
| Overall coverage (%) | **94.8%** |
| Articles with complete coverage | **86/105 (81.9%)** |
